# Combination of *BRCA* deep targeted sequencing and shallow whole genome sequencing to detect homologous recombination deficiency in ovarian cancer

**DOI:** 10.1101/2024.09.10.24313378

**Authors:** Thien-Phuc Hoang Nguyen, Nam HB Tran, Tien Anh Nguyen, My TT Ngo, Anh Duong Doan, Du Quyen Nguyen, Hung Sang Tang, Duy Sinh Nguyen, Cam Tu Nguyen Thi, Thanh Thuy Do Thi, Hoai-Nghia Nguyen, Hoa Giang, Lan N Tu

**Affiliations:** Medical Genetics Institute, Ho Chi Minh city, Vietnam; Gene Solutions, Ho Chi Minh city, Vietnam

**Keywords:** homologous recombination deficiency, ovarian cancer, PARPi, *BRCA1/2*, genomic instability

## Abstract

**Backgrounds:** Assessing homologous recombination deficiency (HRD) has been recommended by clinical guidelines for patients with ovarian cancer (OC) as it predicts sensitivity to poly (ADP-ribose) polymerase inhibitors (PARPi). However, HRD testing is complex and either inaccessible or unaffordable for majority of OC patients in developing countries. Consequently, the prevalence of HRD in OC remains unknown.

**Methods:** We examined HRD status of 77 Vietnamese patients with OC using a new laboratory-developed test (HRD Insight, Gene Solutions). Tumor DNA was extracted from FFPE samples, followed by next-generation sequencing to detect deleterious or suspected deleterious variants in *BRCA1*/*2* genes. Shallow whole genome sequencing was performed to determine the whole Genomic Instability (wGI) score by assessing the presence of large-scale intra-chromosomal copy number alterations.

**Results:** The assay was first benchmarked against commercial HRD kits including TruSight Oncology 500 HRD (Illumina), SOPHiA DDM HRD Solutions (Sophia Genetics) and HRD Focus Panel (AmoyDx), and showed an overall percent agreement of 90.0%, 96.3%, and 96.4% respectively. The successful rate of sequencing was 94.8% (73/77) and the prevalence of HRD in OC patients was 54.8% (40/73). *BRCA* mutations and positive wGI scores were found in 16.4% (12/73) and 47.9% (35/73) of the patients respectively. Among those with wild-type *BRCA1*/*2*, 40.5% of them had positive wGI scores and hence positive HRD. Age at diagnosis was not affected by both *BRCA* and wGI status.

**Conclusions:** HRD Insight assay could accurately and robustly determine the HRD status of ovarian tissue samples, including those with low quality.

## Introduction

Homologous recombination deficiency (HRD) occurs when cells cannot repair DNA double-strand breaks via homologous recombination repair (HRR) pathways. It results in genomic instability and subsequent accumulation of DNA damage and genetic alterations. HRD is found the most prevalent in ovarian and breast cancer, followed by prostate and pancreatic cancer [1]. Particularly in advanced ovarian cancer (OC), HRD is considered a predictive biomarker for poly(ADP-ribose) polymerase inhibitors (PARPi) such as olaparib and niraparib, as these drugs significantly improved progression-free and overall survival of OC patients positive for HRD [2]. Therefore, assessing HRD status has been recommended by both NCCN and ESMO guidelines to identify OC patients that are likely to benefit from PARPi therapy [3, 4].

HRD is best characterized by germline or somatic mutations in *BRCA1* and *BRCA2* genes, and earlier HRD testing was based on identifying the presence of deleterious or suspected deleterious mutations in *BRCA1/2*. However, this approach failed to capture epigenetic changes such as *BRCA1* promoter methylation, mutations in other HRR genes, as well as unidentified non-*BRCA* mechanisms [4]. Therefore, current HRD testing further assesses the presence of genomic “scars”, or the large chromosomal and subchromosomal abnormalities that result from HRD irrespective of the underlying causes. The typical types of abnormalities examined in OC are loss of heterozygosity (LOH), telomeric-allelic imbalance (TAI) and large scale transitions (LST) [4, 5]. The addition of TAI and LST markers has been shown to improve the ability to detect HRD in comparison with the LOH marker alone [5]. In recent years, copy number alterations (CNA) emerged as a new HRD biomarker to detect gain or loss of genomic regions as a result of genomic instability. While detecting LOH and TAI markers requires deep genomic profiling data, CNA signals could be captured by shallow whole genome sequencing (sWGS) data, making it a simplified and cost-effective alternative [6]. Commercial tests utilizing either LOH or CNA markers are both recommended by the ESMO guidelines to determine HRD status for OC patients [7].

In developing countries like Vietnam, HRD testing remains mostly inaccessible and/or unaffordable for OC patients due to high cost and technology complexity [8]. Issues with pathological specimens such as DNA integrity and tumor cellular fraction also pose a significant challenge for test implementation [9, 10]. The lack of HRD testing leads to unknown prevalence of HRD in OC patients as well as the general unfamiliarity among local oncologists with its clinical significance. Together, this situation ultimately limits the access to PARPi treatment in a subset of OC patients.

In this study, we developed the HRD Insight (HRD INSI) test that detects mutation status of *BRCA1/2* genes and evaluates genomic instability using the CNA marker. The workflow was optimized for samples with low quality and high sequencing noise, and the performance was benchmarked against other commercial tests. The first HRD spectrum of Vietnamese patients with OC was also presented.

## Materials and Methods

### Sample collection

For clinical samples, formalin-fixed paraffin-embedded (FFPE) tumor samples of newly diagnosed OC patients were obtained from the Medical Genetics Institute, Ho Chi Minh city, Vietnam and stored for less than 2 years.

The study was conducted in accordance with the ethical principles of the Declaration of Helsinki. Ethical approval was obtained from the institutional ethics committee of the Medical Genetic Institute, Ho Chi Minh City (approval number 03/2024/CT-VDTYH). All samples and their genomic data were de-identified and aggregated for analysis.

### *BRCA1/2* deep targeted sequencing and shallow WGS

Genomic DNA was already isolated from FFPE samples and subjected to a standard library preparation protocol as previously described [11]. A minimum input of 8 ng gDNA and 150 ng of library yield was required for the whole workflow. Part of the DNA libraries were pooled and hybridized with a probe panel consisting of *BRCA1* and *BRCA2* genes (Integrated DNA Technologies IDT, USA). Deep sequencing of enriched libraries was performed on the DNBSEQ-G400 sequencer (MGI, China) using 2×100 bp paired-end sequencing and achieving an average depth of 100X per sample. The rest of the DNA libraries were subjected to sWGS on the DNBSEQ-G400 sequencer (MGI, China) using 2×100 bp paired-end sequencing and achieving an average depth of 1-2X per sample. For samples that failed to meet the minimum library yield or the bioinformatics quality control (QC) criteria described below, a modified protocol was applied to prepare DNA libraries using the NEBNext UltraShear FFPE DNA Library Prep Kit (New England Biolabs, USA) according to the manufacturer’s instructions.

### Bioinformatics analysis

For *BRCA1/2* mutation status, samples passed QC if the mean depth of the *BRCA1/2* region was greater than 40X and the percent coverage at 40X was equal to or exceeded 90%. Single nucleotide variant (SNV) and short insertion-deletion (indel) variant calling was performed using the DRAGEN™ Bio-IT Platform (v3.10). The effect and population frequency of variants were predicted by VEP (version 105) [12] and annotated against the dbSNP [13], ClinVar [14], and COSMIC [15] databases. Variants with a VAF < 5% and high population frequency (>0.1%) were excluded from the analysis. For large genomic rearrangements (LGRs) of *BRCA*, they were called using the DRAGEN™ Structural Variant Caller pipeline, which leverages paired and split-read mapping information [16]. LGRs that passed the QC metrics in the structural variant caller were included for post-analysis. All variants were classified, interpreted and reported according to the guidelines of the Association for Molecular Pathology, American Society of Clinical Oncology, and College of American Pathologists [17]. Samples harboring any deleterious or suspected deleterious variants were considered positive for *BRCA1/2* mutation status.

For genomic instability status, samples passed QC if the mean depth was greater than 1X and a percent coverage at 1X was equal to or exceeded 40%. Tumor fraction (TF) of the samples was determined by ichorCNA [18]. Copy number alteration (CNA) profiles were constructed from sWGS data using QDNAseq (v1.39) [19] based on read counts in fixed window sizes. The results were then analyzed by shallowHRD (v1.13) [20] to detect large-scale genomic alterations (LGAs), which are markers of genomic instability. The pipeline was further modified to enhance its performance to detect genomic instability. During the bin annotation step performed by QDNAseq, only properly paired reads that successfully aligned to the reference genome were included, and read counts were normalized using larger window sizes to minimize artifacts from FFPE samples. The median and standard deviation across all defined segments were calculated to estimate noise signals across the genome. In the shallowHRD step, factors such as noise from FFPE samples and TF were considered to establish minimal CNA cut-off values, ensuring harmonization in CNA event calculations. From CNA events, LGAs were defined as chromosome arm breaks between adjacent genomic segments (less than 3 Mb apart) of more than 10 Mb. The number of LGA events was then scaled, with a score of 0 serving as the cut-off for the whole-genome instability (wGI) score. Samples with a wGI score above 0 were classified as positive for genomic instability.

### Benchmarking analysis

Our assay was benchmarked against commercial orthogonal tests for a subset of clinical samples. The commercial tests were the HRD Focus Panel (AmoyDx, China), SOPHiA DDM HRD Solutions (SOPHiA Genetics, France), and TruSight Oncology 500 HRD (TSO500 HRD) (Illumina, USA). Library preparation, target enrichment and sequencing for FFPE samples were performed according to the each manufacturer’s protocols. Data were analyzed using the respective manufacturer’s platform to determine the QC and HRD status.

### Analysis of DNA integrity and tumor fraction

For DNA integrity number (DIN) assessment, 20 ng of DNA was used for automated electrophoresis using the Genomic DNA ScreenTape reagent (Agilent, USA).

For in-silico TF simulation, FFPE with TF of ≥ 60% (as determined by both our pipeline and pathological assessment) and matching white blood cell (WBC) samples were subjected to sWGS at the depth of ∼1X. Unique reads from processed BAM files of paired FFPE and WBC were extracted for in-silico simulation. Tumor reads were mixed with WBC reads to create simulated TF ranging from 60% to 10% with a 10% decrement. The simulated samples were then assessed for wGI score using sWGS data and TF estimation by ichorCNA.

### Statistical analysis

Sensitivity, specificity, positive percent agreement (PPA), and negative percent agreement (NPA) were calculated using RStudio. Correlation analysis between our test and the reference kits was performed by calculating the Pearson correlation coefficient using the R v4.3.0 stats package. Student’s t-test was performed using Prism 10 version 10.2.3 (GraphPad Software LLC, USA) with significant p value defined as below 0.05.

## Results

### Assay and study workflow

In the HRD INSI assay workflow, genomic DNA was extracted from FFPE ovarian tissue samples and whole genome DNA library was prepared (Figure 1A). Part of the DNA library was hybridized with a gene panel consisting of *BRCA1* and *BRCA2* genes to identify deleterious or suspected deleterious variants. The rest of the DNA library was subjected to sWGS (1-2X) to identify all CNA events, which were converted to a whole genomic instability (wGI) score. CNA was used as a marker for genomic instability as it captured large genomic alterations that could also reflect LOH, TAI and LST occurrence (Figure S1) [21]. The full pipeline of HRD analysis (Figure S2A) illustrated data pre-processing steps including adapter removal, low-quality base trimming, reference genome mapping and QC; followed by analysis for *BRCA1/2* mutations and wGI score as described in the Methods. Representative outputs from the pipeline for samples with positive- and negative-wGI scores (Figure S2B) included in-silico TF estimation, noise index representing the median fluctuation of bins within each segment across the whole genome, CNA cut-off values that were threshold baselines used to identify LGA events, and final wGI score. Overall, a tumor was determined as HRD-positive when either *BRCA1/2* mutation status or wGI score was positive.

**Figure 1.**
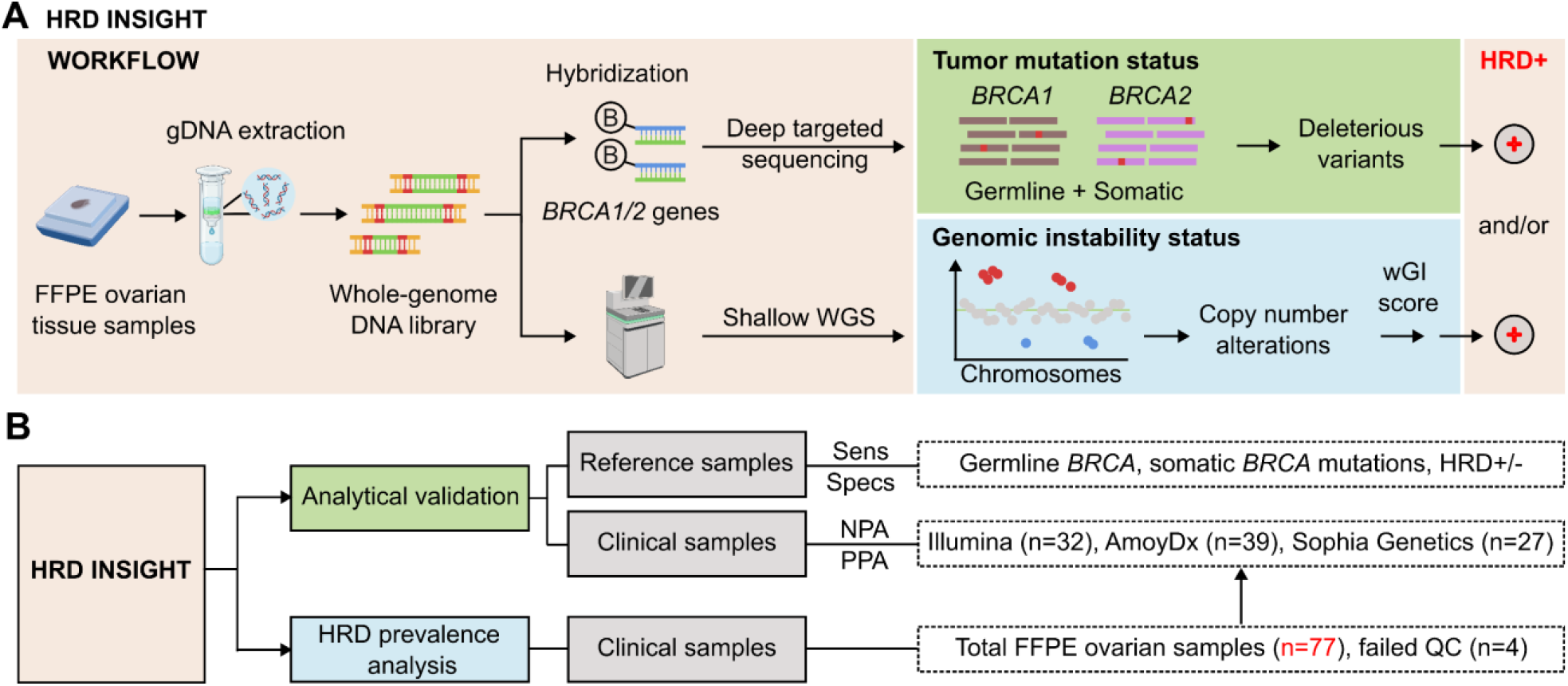
Study design and workflow of HRD INSIGHT assay. **(A)** Assay workflow: genomic DNA was first extracted from FFPE ovarian tissue samples. DNA library was prepared and then hybridized with a gene panel consisting of *BRCA1* and *BRCA2* genes to identify deleterious or suspected deleterious variants. The DNA library was also subjected to shallow whole genome sequencing (WGS) to identify copy number alteration events, which were converted to a whole genomic instability (wGI) score to determine genomic instability status. A sample was determined as HRD-positive when either *BRCA1/2* mutation status or wGI score was positive. (Created with Biorender.com). **(B)** The study design consisted of analytical validation and HRD prevalence analysis. For analytical validation, reference standards were used to determine the sensitivity and specificity of the HRD INSIGHT assay; clinical samples were used to determine the negative- and positive-performance agreement (NPA and PPA) with commercial HRD kits. The HRD prevalence analysis utilized 77 FFPE ovarian samples, 4 of which failed quality control (QC).H

Analytical validation was first performed using reference standards to determine the sensitivity, specificity and reproducibility of the HRD INSI assay to detect both germline and somatic *BRCA* mutations, as well as genomic instability (Figure 1B). Clinical samples of FFPE ovarian tissue samples were then used to further evaluate PPA and NPA of the HRD INSI with commercial HRD kits. Subsequently, the HRD status of 77 ovarian samples from Vietnamese patients was examined (Figure 1B).

### Development and analytical validation of HRD INSIGHT

We used the reference standards BRCA Germline I (Horizon, USA) that had 13 verified germline variants (10 SNVs and 3 small Indels) and OncoSpan FFPE (Horizon, USA) that had 6 verified somatic variants (4 SNVs and 2 small Indels) in *BRCA1* and *BRCA2* genes. Sensitivity of germline mutation detection was estimated from 15 replicates: 5 replicates per run for 3 independent runs, while sensitivity of somatic mutation detection was estimated from 11 replicates: 2-3 replicates per run for 4 independent runs. The reference standards, BRCA Germline I (Horizon, USA) and Tru-Q 0 (Horizon, USA), reported to have no certain BRCA germline and somatic variants respectively, were used to determine specificity of the assay. For germline variants, the HRD INSI assay detected both SNV and small Indel variants with sensitivity and specificity of >99% (Figure 2A). For somatic variants, sensitivity was estimated to be >99% for variants with VAF ≥ 5% and specificity was >99%. There was high correlation R^2^ = 0.99 between the expected VAFs and VAFs determined by the HRD INSI assay (Figure 2A). For *BRCA* germline LGRs, the HRD INSI assay could detect 67% (8/11) of the mutations in the Seraseq® FFPE BRCA1/2 LGR Reference Material (SeraCare, USA) (Figure 2B). The 3 LGRs that could not be detected were deletions with a size range of 50-200 bp, while the other LGRs of similar or larger sizes were still detected (Figure 2B).

**Figure 2.**
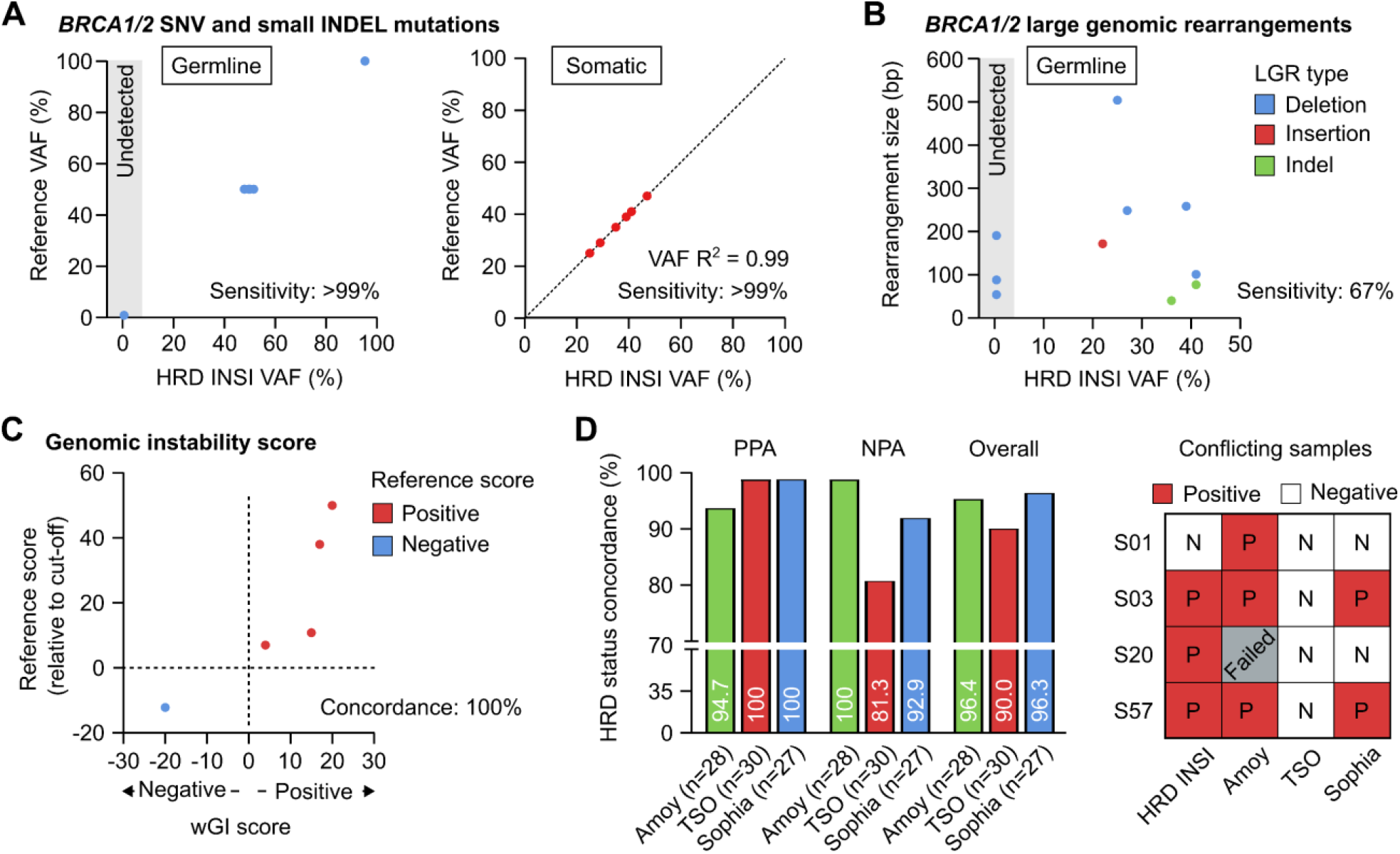
Analytical validation of the HRD INSIGHT assay. Sensitivity of the HRD INSIGHT assay to detect **(A)** germline and somatic SNV and small Indel mutations, **(B)** germline large genomic rearrangements (LGRs) of BRCA1/2 genes in reference standards. **(C)** Concordance of the HRD INSIGHT assay to determine the genomic instability score and status of the reference standards. **(D)** Positive-, negative- and overall-percent agreement (NPA, PPA, Overall) of the HRD INSIGHT assay with the HRD Focus Panel (AmoyDx), TruSight Oncology TSO 500 HRD (Illumina) and SOPHiA DDM HRD Solutions (Sophia Genetics) to determine HRD status of clinical FFPE ovarian samples. For 4 conflicting samples, the results of individual assays were shown.

For genomic instability score, 5 reference standards used to validate our wGI score algorithm were Seraseq*®* FFPE HRD High-Positive Reference Material, Seraseq*®* FFPE HRD Low-Positive Reference Material (Seracare, USA), GIInger™ Positive, GIInger™ Negative (Sophia Genetics, USA) and HRD Positive control (AmoyDx, China) (Figure 2C). The HRD INSI showed concordance of 100% to determine both positive and negative reference standards. Moreover, there was a trend that the samples with higher reference scores also received higher wGI scores (Figure 2C).

We then compared performance of the HRD INSI assay with the HRD Focus Panel (AmoyDx), TruSight Oncology TSO 500 HRD (Illumina) and SOPHiA DDM HRD Solutions (Sophia Genetics) using 28-, 30-, and 27-FFPE ovarian samples respectively. The overall percent agreement between the HRD INSI and AmoyDx, Illumina TSO and Sophia kits were 96.4%, 90.0% and 96.3% respectively (Figure 2D). While the PPA and NPA among all assays were mostly above 90%, the HRD INSI assay shared the lowest PPA with the AmoyDx kit (94.7%) and the lowest NPA with the Illumina TSO kit (81.3%). This was because the AmoyDx kit classified more *BRCA* mutations as “deleterious” compared to all other assays (Figure S3A); and the Illumina TSO kit found negative genomic instability score in more samples than the others (Figure S3B). Among the 4 conflicting samples, the HRD status of 3 samples S01, S03 and S57 as determined by the HRD INSI assay agreed with 2 out of 3 commercial kits (Figure 2D). Sample S20 had low DNA quality and failed the AmoyDx test, it was positive with a borderline wGI score of 0 by HRD INSI assay and was negative by the Illumina TSO and Sophia kits (Figure 2D).

The HRD INSI assay had a successful rate of 94.8%, comparable with that of the Illumina TSO (93.8%) and Sophia (96.2%) tests and higher than that of the AmoyDx test (71.8%) (Figure 3A). When a subset of 42 samples was analyzed, the quality of DNA isolated from FFPE samples was found as the crucial factor to determine successful analysis. DNA quality was reflected by DNA integrity number (DIN) of the sample input and the noise index after sequencing (Figure 3B). Most of the samples with noise index level above 0.10 failed the AmoyDx test and those with DIN lower than 2 also had a high failure rate. In our workflow, we implemented modifications to the original shallowHRD pipeline, including an increase of window size in the bin annotation step from 50 kbp to 500 kbp to reduce artifact signals from degraded FFPE samples, and incorporation of noise index and in-silico TF values to better harmonize CNA event calculation. The R^2^ value between our wGI score with TSO GIS score was significantly improved with the modified pipeline (R^2^ = 0.83) compared to the original pipeline (R^2^ = 0.63) (Figure 3C). Apart from bioinformatics optimization, a modified wet lab protocol for DNA library preparation using the NEBNext UltraShear kit was also applied for samples with low DNA quality. Due to the limited number of samples that failed QC in this cohort, we used a different set of in-house FFPE DNA samples (n = 20) to demonstrate the effectiveness of this modification. Using same DNA input, the DNA library yield, QC measures for *BRCA1/2* targeted sequencing were significantly improved for the modified protocol compared to the standard protocol (Figure S4A-B). Specifically, 0% (0/20) of the samples prepared by the standard protocol achieved 90% coverage at 40X for *BRCA1/2* and hence failed QC, while 85% (17/20) of them passed QC when switched to the modified protocol (Figure S4B). QC for sWGS data were not different between the two protocols (Figure S4C).

**Figure 3.**
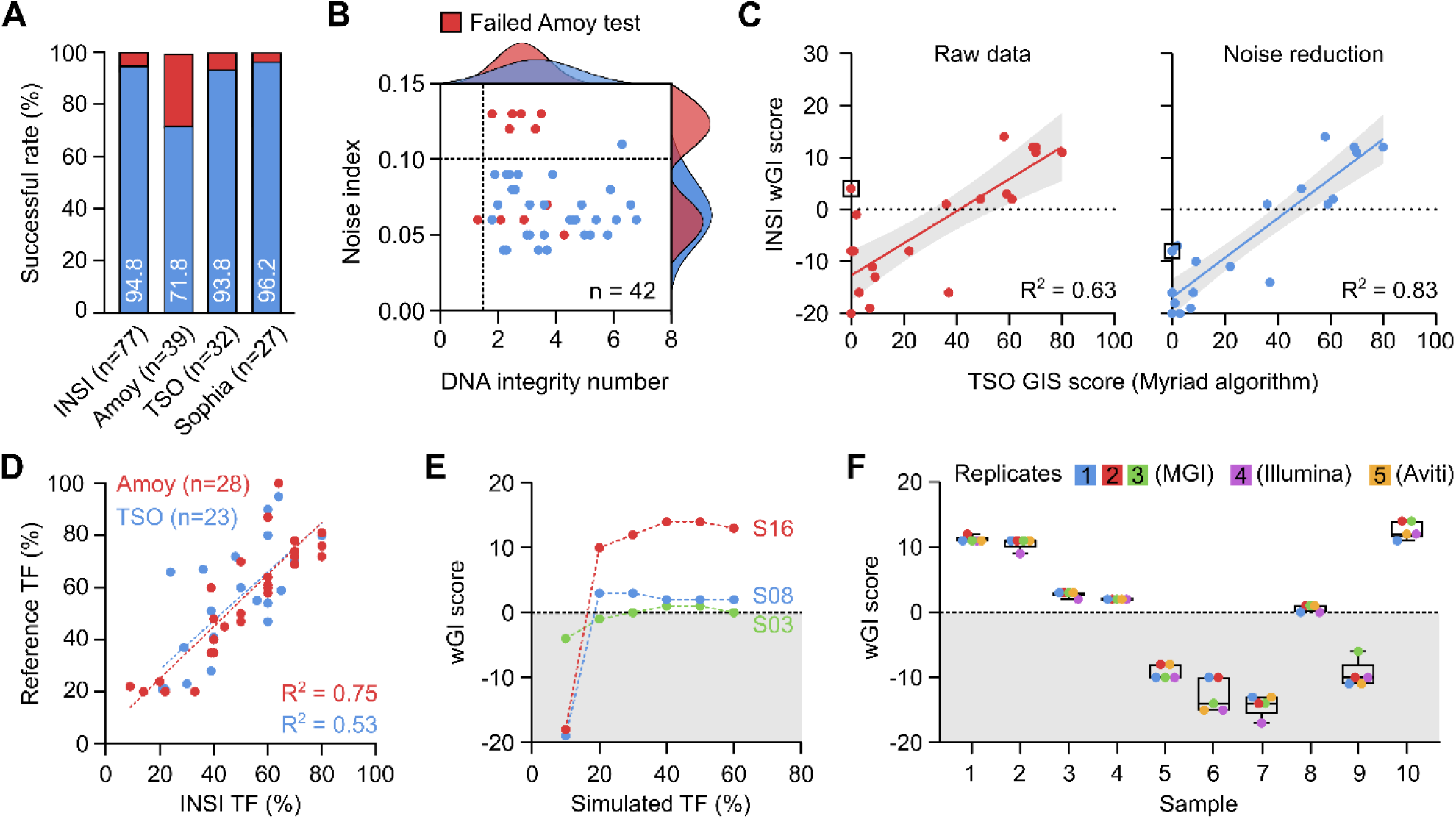
Quality management of the HRD INSIGHT assay. **(A)** The successful rate of each assay to determine HRD status of clinical samples. **(B)** Distribution of noise index and DNA integrity number among samples that failed AmoyDx test. **(C)** Noise reduction algorithm improved the correlation of wGI score with TSO genomic instability (GIS) score. **(D)** Tumor fraction (TF) was determined in-silico and compared with AmoyDx and TSO kits. **(E)** In-silico simulation of TF serial dilution at different levels showed stable wGI scores at a minimum TF of 30%. **(F)** wGI score measurement was robust and reproducible among different runs across different sequencing platforms.

Besides DNA quality, TF of a FFPE sample was a known factor to cause false negative result. Our pipeline estimated in-silico TF that showed a high correlation with the TF estimated by AmoyDx and Illumina TSO tests (Figure 3D). When we performed in-silico simulation of TF serial dilution to different levels, the wGI score remained stable at TF of 20% for samples S16 and S08, but for sample S03 with a borderline wGI score, TF of at least 30% was required to ensure accuracy (Figure 3E). Therefore, the requirement of TF was set at 30% for our assay. The high repeatability and reproducibility of the HRD INSI assay using clinical samples was demonstrated among different runs and across different NGS platforms, including samples with borderline wGI scores (Figure 3F). The overall technical performance and quality control measures of the HRD INSI assay are listed in Table 1.

**Table 1.** Overall performance and quality control parameters of HRD INSIGHT.

| TECHNICAL PERFORMANCE |  |  |
| --- | --- | --- |
|  | Sensitivity | Specificity |
| BRCA1/2 germline variants |  |  |
| SNV | >99% | >99% |
| INDEL (<50 bp) | >99% | >99% |
| BRCA1/2 somatic variants (at VAF ≥ 5%) |  |  |
| SNV | >99% | >99% |
| INDEL (<50 bp) | >99% | >99% |
| Genomic instability status | >99% | >99% |
| HRD status of clinical samples (benchmarked against SOPHiA DDM™ HRD Solution*) |  |  |
| Positive percent agreement | 100% |  |
| Negative percent agreement | 92.9% |  |
| Overall percent agreement | 96.3% |  |
| QUALITY CONTROL |  |  |
|  | Shallow WGS | Targeted sequencing |
| FFPE samples |  |  |
| Tumor fraction | ≥30% | ≥30% |
| DNA integrity number | ≥1.5 | ≥1.5 |
| Minimum DNA input | ≥8 ng | ≥8 ng |
| Sequencing Data (for MGI, Illumina and Element Biosciences platforms) |  |  |
| Mean depth | ≥1X | ≥40X |
| Mean coverage | ≥40% at 1X | ≥90% at 40X |
| Read length | 100bp x 2, 150bp x 2 | 100bp x 2, 150bp x 2 |

\* Recommended by ESMO for HRD testing.
SNV: single nucleotide variation; Indel: insertion/deletion; VAF: variant allele frequency.

### Prevalence of HRD in Vietnamese patients with ovarian cancer

Using the HRD INSI test, we analyzed the HRD status of 77 FFPE tissue samples from Vietnamese patients with OC. The average age at diagnosis was 54 years old and majority(84.4%) of the patients had advanced OC at stage III-IV. Most cases (64.9%) did not provide the histological subtypes (Table S1).

Out of 77 samples, 4 samples failed QC due to either insufficient DNA amount or low DNA quality. Among the 73 samples with successful sequencing, the prevalence of HRD was 54.8% (40/73). A deleterious or suspected deleterious *BRCA* mutation was identified in 16.4% (12/73) patients while wGI score was positive in 47.9% (35/73) patients (Figure 4A). We compared these positive rates with other Asian and Caucasian populations [10, 22-27] and observed that the rates of *BRCA* mutation and overall HRD positivity in other Asian studies were mostly above 20% and 60% respectively, slightly higher than those in our study. The positive rate of genomic instability score and overall HRD status in our study, similar to other Asian populations, appeared higher than those of the Caucasian populations, except the Russian (Figure 4A). Among HRD-positive samples, we observed a lower proportion of patients having both *BRCA* mutation and positive genomic instability score (22.5%) and a higher proportion having a positive score alone (70.0%), compared to other studies (Figure 4B). Moreover, the wGI scores were significantly higher in patients with mutated *BRCA1/2* compared to those with wild-type *BRCA1/2*. The proportion of patients with a positive wGI score were 40.5% and 75.0% in those with wild-type and mutated *BRCA1/2* respectively (Figure 4C). There was no difference in the age at diagnosis for patients with different wGI and BRCA mutation status (Figure 4D). Finally, the mutations found in *BRCA1/2* distributed along the genes with no obvious hotspots (Figure 4E). The details of wGI and *BRCA* mutation status of each patient are listed in Table S2.

**Figure 4.**
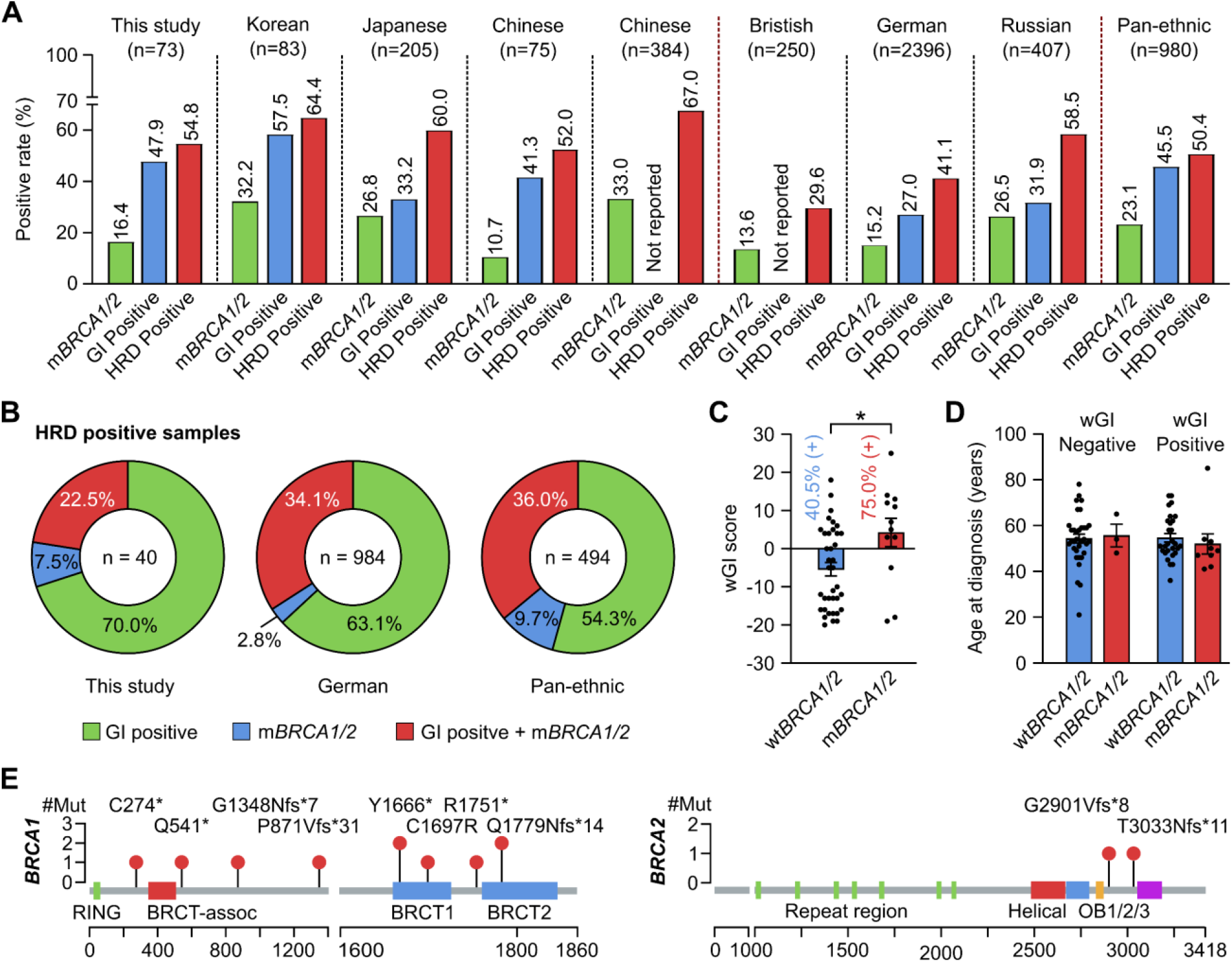
Analysis of HRD status in Vietnamese patients with ovarian cancer. **(A)** The positive rates of HRD, mutated BRCA1/2 (mBRCA1/2) and genomic instability (GI) status in this study were compared with other patient cohorts. **(B)** Among HRD-positive samples, the proportion of those having both GI-positive and mBRCA1/2 was 22.5% and compared with other patient cohorts. **(C)** wGI score was significantly higher in samples with mBRCA1/2 compared to wild-type BRCA1/2 (wtBRCA1/2). *p < 0.05 by Student’s t-test. **(D)** Age at diagnosis was not affected by the wGI or BRCA1/2 mutation status. **(E)** Distribution of mutations identified in our cohort along the BRCA1 and BRCA2 proteins.

## Discussion

In this study, we developed and validated a HRD assessment workflow that was based on *BRCA1/2* mutation testing and CNA-based wGI score. Although several tests such as FoundationOne and MyChoice^®^CDx measure LOH, TAI and LST for genomic instability, there are certain disadvantages associated with these complex biomarkers. They tend to require deep genomic profiling data, harder to acquire in samples with low tumor content or low DNA quality, and the optimal cut-off is not consistent among different types of cancer [1, 6, 28, 29]. On the other hand, CNA is an inherent signature in multiple types of cancer and could be assessed by a wider range of techniques, even in samples with low tumor cellularity, making it a cost-effective and feasible approach. Moreover, CNA has been utilized in the ESMO-recommended test kit SOPHiA DDM™ HRD Solution (Sophia Genetics, USA) [7] and well validated as a biomarker of HRD, not only in OC but also other cancer types [6, 28, 29]. For those reasons, we selected CNA profiling by sWGS as a practical and affordable solution for HRD testing in developing countries.

Our analytical validation and benchmarking analysis evidently showed that the HRD INSI assay had good performance and high concordance with orthogonal test kits. Particularly in comparison with the Sophia Genetics test that also uses CNA marker, our overall percent agreement (96.3%), PPA (100%) and NPA (92.9%) were higher than those of previously developed tests, such as Jun Kang *et al* that reported 91.1%, 91.5% and 90% respectively [27]. As expected, discordance was still observed among different HRD test kits in some cases. The results of HRD INSI were in agreement with 2 out of 3 commercial tests in most of conflicting cases, indicating the reliability and accuracy of our test. Other observations including the higher HRD positive rate using AmoyDx test and discordance in classification of *BRCA* variants were similar to previous reports [30-32]. Furthermore, the successful rate of our assay achieved 94.2%, higher than AmoyDx and comparable with Sophia and Illumina TSO kits. Low quality of DNA samples was the main factor for failure. Our modified wet lab protocol and optimized bioinformatics pipeline were shown to improve the failure rate and assay performance. Besides DNA quality, TF has been undoubtedly demonstrated as a significant factor influencing accuracy of HRD testing. TF below 30%-40% as determined by pathological assessment could lead to a false negative result [10, 33]. Since information of tumor cellularity of tissue specimens is often lacking in local pathology practice, the inclusion of in-silico TF estimation in our pipeline is helpful to inform doctors of a potentially inaccurate result.

Since HRD testing is not yet widely available in Vietnam, the prevalence and spectrum of HRD is currently unknown in the Vietnamese patients with OC. *BRCA* mutational analysis, however, has been performed for both germline and somatic variants. Vu *et al* previously reported a *BRCA1* mutation rate at 7.9% (8/101) in the Vietnamese OC patients with mixed subtypes and found no *BRCA2* mutation in the cohort [34]; while Chu *et al* reported a mutation rate of 15.2% (5/33) in both *BRCA1* and *BRCA2* genes [35]. In this study, we identified both *BRCA1* and *BRCA2* mutations at the overall frequency of 16.4% (12/73), albeit lower than other Asian cohorts but higher than those studies in the Vietnamese patients. Such difference is likely atributed to the proportion of each histological subtype in the studied cohorts, as it has been shown that serous carcinoma had the highest rate of *BRCA* mutations, endometrioid carcinoma had a very low mutation rate while the mucinous and clear cell subtypes did not have any *BRCA* mutations [34]. The histological subtype information of our cohort unfortunately was not available in most cases, but it was likely to be of mixed subtypes, similar to the two Vietnamese cohorts above and different from other Asian studies that were more enriched with serous ovarian carcinoma. The spectrum of *BRCA* mutations identified in this study included all common mutations previously reported such as p.Gln541Ter, p.Arg1751Ter, and p.Gln1779AsnfsTer14 [34-36]. Besides *BRCA* status, the lack of subtype information could also affect the interpretation of HRD prevalence as the rate of HRD has been shown to be the highest in high-grade serous ovarian carcinoma (HGSOC) compared to all other subtypes [37]. Hence, the HRD prevalence estimated from our mixed cohort could be an underestimation for HGSOC patients, explaining our lower HRD rate compared to other Asian cohorts [24, 25, 27]. Furthermore, we found that patients carrying *BRCA1/2* mutations had a significantly higher genomic instability score than those with wild-type *BRCA1/2*, which was documented previously [24]. It should be emphasized that 40.5% of patients without any *BRCA* mutations still presented with genomic instability established by positive wGI scores. This result strongly advocates for comprehensive HRD assessment instead of *BRCA1/2* testing alone in OC patients so that no patients miss their opportunity for PARPi treatment.

Our study was limited by the small sample size as well as the lack of subtype information for more thorough analysis. Moreover, growing evidence has corroborated the involvement of other HRR genes beyond *BRCA* or *BRCA* variant of unknown significance in HRD [1, 4]. While a larger gene panel consisting of more HRR genes is now recommended for HRD research, the pathogenicity of these variants remains unclear and current clinical guidelines still exclude them in HRD result interpretation, which might be changed in the near future. Besides the extension of genes and variants, several studies also found that HRD status could predict response to platinum-based chemotherapy in triple negative breast cancer [38] or immuno-neoadjuvant therapy in non-small cell lung cancer [39]. Therefore, future CNA-based HRD assessment could be amenable to other cancer types for other indications beyond PARPi [28].

## Data Availability

All data produced in the present work are contained in the supplementary

## Author contributions

Thien-Phuc Hoang Nguyen, Tien Anh Nguyen, Anh Duong Doan, Hoa Giang performed bioinformatics and statistical analysis. Nam HB Tran, My TT Ngo, Cam Tu Nguyen Thi processed standard and clinical samples for sequencing. Thanh Thuy Do Thi, Du Quyen Nguyen, Hung Sang Tang, Duy Sinh Nguyen, Hoai-Nghia Nguyen performed clinical analysis. Thien-Phuc Hoang Nguyen, Nam HB Tran, Lan N Tu designed experiments, analyzed data and wrote the manuscript.

