## Supplementary table and figure for "Combination of *BRCA* deep targeted sequencing and shallow whole genome sequencing to detect homologous recombination deficiency in ovarian cancer"

**Table S1. Patient demographics**

| **Characteristic** | 77 (100%) |
|---|---|
| **Age at diagnosis (range), years** | 54 (21 – 85) |
| >50 | 47 (61.0%) |
| ≤50 | 30 (39%) |
| **Subtype (N, %)** | |
| High-grade serous | 22 (28.6%) |
| Endometrioid | 4 (5.2%) |
| Mucinous | 1 (1.3%) |
| Unknown | 50 (64.9%) |
| **Stage (N, %)** | |
| I | 6 (7.8%) |
| II | 6 (7.8%) |
| III-IV | 65 (84.4%) |

**Table S2. *BRCA* mutation status and wGI score of ovarian cancer patients**

| **ID** | **BRCA1/2 clinically significant mutation** | | | **Instability score** | | **HRD status** |
|---|---|---|---|---|---|---|
| | Mutation | VAF | Conclusion | wGI | Conclusion | |
| S01 | ND | | NEG | -16 | NEG | **NEG** |
| S02 | ND | | NEG | 3 | NEG | **NEG** |
| S03 | ND | | NEG | 1 | POS | **POS** |
| S04 | ND | | NEG | 3 | POS | **POS** |
| S05 | ND | | NEG | -10 | NEG | **NEG** |
| S06 | ND | | NEG | -7 | NEG | **NEG** |
| S07 | ND | | NEG | 3 | NEG | **NEG** |
| S08 | ND | | NEG | 2 | POS | **POS** |
| S09 | *BRCA2:* 3033Nfs*11 | 38% | POS | -18 | NEG | **POS** |
| S10 | ND | | NEG | -12 | NEG | **NEG** |
| S11 | ND | | NEG | -16 | NEG | **NEG** |
| S12 | ND | | NEG | -19 | NEG | **NEG** |
| S13 | ND | | NEG | 11 | POS | **POS** |
| S14 | *BRCA1:*Q1779Nfs*14 | 70% | POS | 11 | POS | **POS** |
| S15 | ND | | NEG | 12 | POS | **POS** |
| S16 | ND | | NEG | 14 | POS | **POS** |
| S17 | ND | | NEG | -13 | NEG | **NEG** |
| S18 | ND | | NEG | -17 | NEG | **NEG** |
| S19 | ND | | NEG | -18 | NEG | **NEG** |
| S20 | ND | | NEG | 0 | POS | **POS** |
| S21 | ND | | NEG | 1 | POS | **POS** |
| S22 | *BRCA1:*R1751* | 64.1% | POS | 14 | POS | **POS** |
| S23 | ND | | NEG | 5 | POS | **POS** |
| S24 | *BRCA1:*P871Vfs*31 | 54% | POS | -5 | NEG | **POS** |
| S25 | ND | | NEG | -5 | NEG | **NEG** |
| S26 | ND | | NEG | 10 | POS | **POS** |
| S27 | ND | | NEG | 5 | POS | **POS** |
| S28 | ND | | NEG | -11 | NEG | **NEG** |
| S29 | ND | | NEG | 11 | POS | **POS** |
| S30 | ND | | NEG | 1 | POS | **POS** |
| S31 | *BRCA1:*Q1779Nfs*14 | 79% | POS | 12 | POS | **POS** |
| S32 | ND | | NEG | -5 | NEG | **NEG** |
| S33 | ND | | NEG | 14 | POS | **POS** |
| S34 | *BRCA1:*C1697R | 84.2% | POS | 13 | POS | **POS** |
| S35 | ND | | NEG | -17 | NEG | **NEG** |
| S36 | ND | | NEG | 0 | POS | **POS** |
| S37 | ND | | NEG | 6 | POS | **POS** |
| S38 | *BRCA1*:Y1666* | 65.4% | POS | 3 | POS | **POS** |

| | | | | | | |
|---|---|---|---|---|---|---|
| S39 | ND | | NEG | -17 | NEG | **NEG** |
| S40 | ND | | NEG | 4 | POS | **POS** |
| S41 | ND | | NEG | -18 | NEG | **NEG** |
| S42 | ND | | NEG | -11 | NEG | **NEG** |
| S43 | ND | | NEG | -13 | NEG | **NEG** |
| S44 | ND | | NEG | -13 | NEG | **NEG** |
| S45 | ND | | NEG | -5 | NEG | **NEG** |
| S46 | ND | | NEG | -19 | NEG | **NEG** |
| S47 | ND | | NEG | -10 | NEG | **NEG** |
| S48 | ND | | NEG | -17 | NEG | **NEG** |
| S49 | *BRCA1:*C274* | 82.7% | POS | 25 | POS | **POS** |
| S50 | ND | | NEG | -16 | NEG | **NEG** |
| S51 | ND | | NEG | 0 | POS | **POS** |
| S52 | ND | | NEG | 4 | POS | **POS** |
| S53 | ND | | NEG | -12 | NEG | **NEG** |
| S54 | ND | | NEG | -3 | NEG | **NEG** |
| S55 | ND | | NEG | -13 | NEG | **NEG** |
| S56 | ND | | NEG | 18 | POS | **POS** |
| S57 | ND | | NEG | 5 | POS | **POS** |
| S58 | ND | | NEG | 8 | POS | **POS** |
| S59 | ND | | NEG | -12 | NEG | **POS** |
| S60 | *BRCA2*:G2901Vfs*8 | 50.0% | POS | -19 | NEG | **POS** |
| S61 | ND | | NEG | 5 | POS | **POS** |
| S62 | ND | | NEG | -16 | NEG | **POS** |
| S63 | ND | | NEG | 0 | POS | **POS** |
| S64 | ND | | NEG | -20 | NEG | **POS** |
| S65 | *BRCA1:*G1348Nfs*7 | 62.7% | POS | 5 | POS | **POS** |
| S66 | ND | | NEG | 4 | POS | **POS** |
| S67 | ND | | NEG | 10 | POS | **POS** |
| S68 | ND | | NEG | 4 | POS | **POS** |
| S69 | ND | | NEG | -16 | NEG | **NEG** |
| S70 | ND | | NEG | -13 | NEG | **NEG** |
| S71 | ND | | NEG | -19 | NEG | **NEG** |
| S72 | *BRCA1:*Q541* | 80.9% | POS | 3 | POS | **POS** |
| S73 | *BRCA1:*Y1666* | 63.6% | POS | 7 | POS | **POS** |
| S74 | ND | | NEG | 7 | POS | **POS** |

*Abbreviations: ND: not detected, NEG: negative, POS: positive, VAF: variant allele frequency*

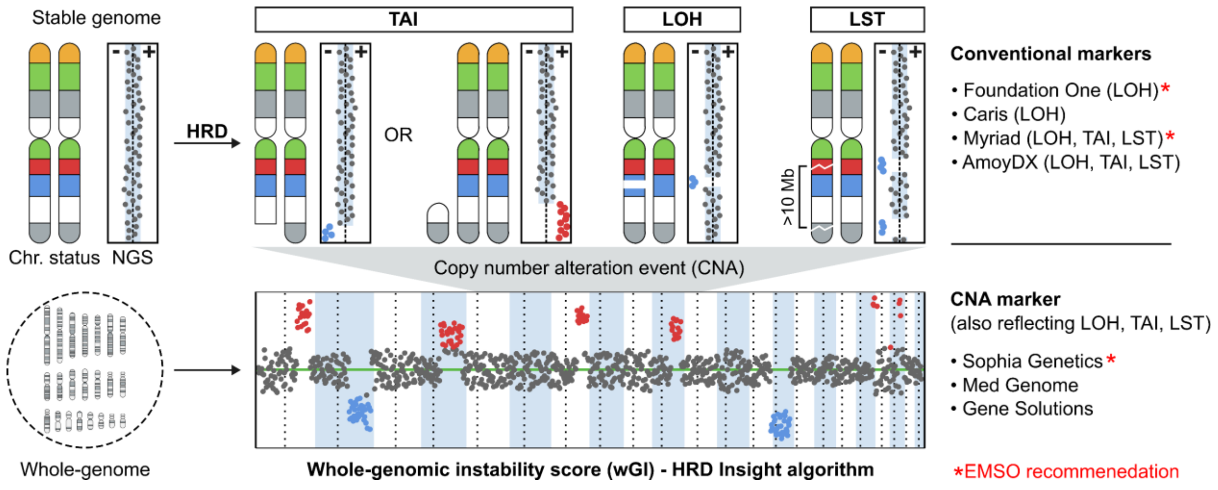


**Figure S1. Biomarkers for homologous recombination repair deficiency (HRD).** HRD in the tumor genome could result in chromosomal and subchromosomal abnormalities such as loss of heterozygosity (LOH), large-scale state transitions (LST) and telomeric allelic imbalance (TAI). Conventional markers of HRD including LOH, TAI, LST are examined in commercial HRD test kits from Foundation One, Myriad and AmoyDx. Copy number alterations (CNA) is a new biomarker that captures large genomic rearrangement events and also reflects LOH, TAI and LST occurrence. It has been examined in commercial HRD kits from Sophia Genetics, Med Genome and Gene Solutions.

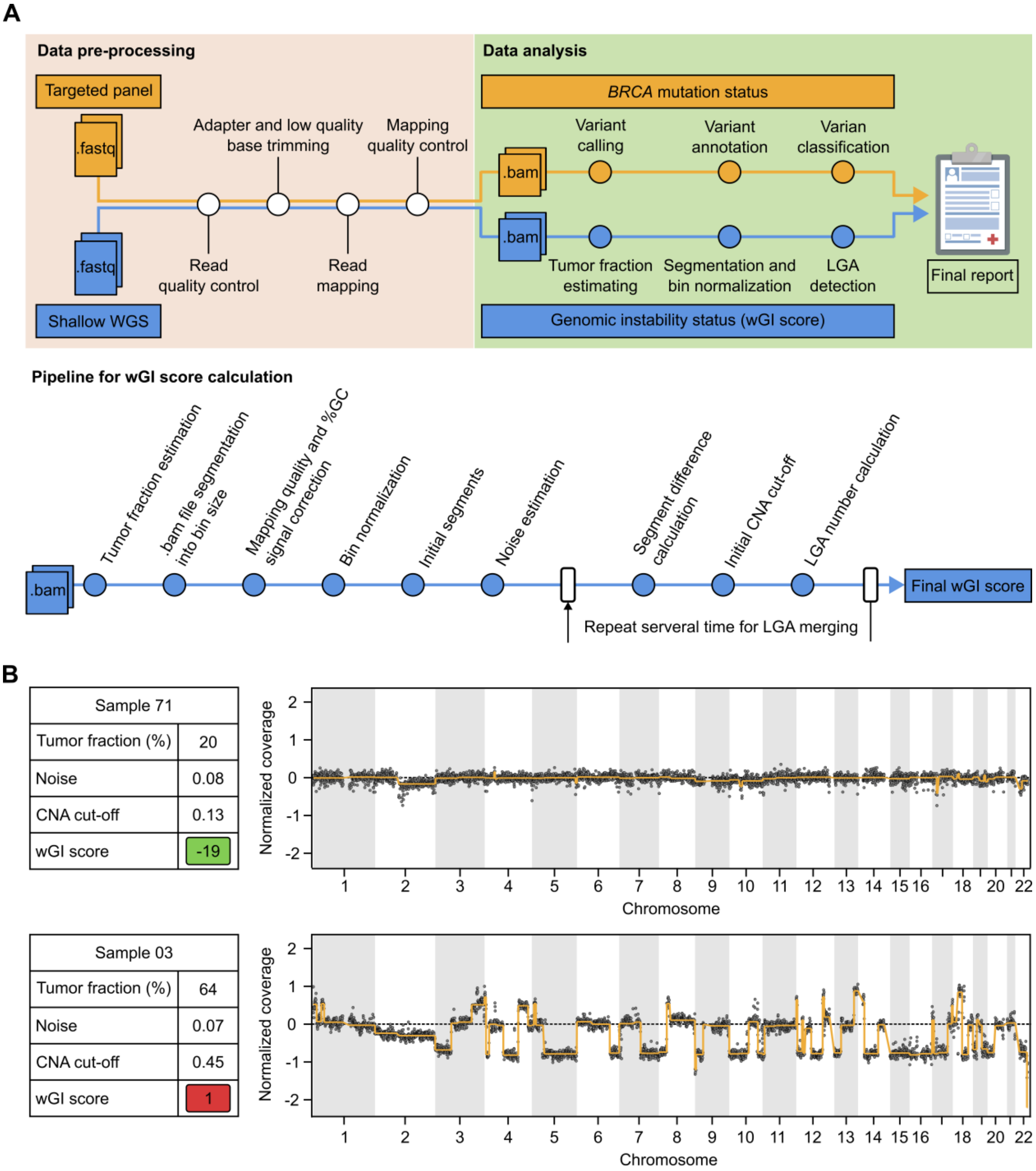


**Figure S2. Data analysis workflow of the HRD INSIGHT assay. (A)** The full pipeline illustrated data pre-processing steps including adapter removal, low-quality base trimming, reference genome mapping and QC; followed by analysis for *BRCA1/2* mutations and wGI score as described in the Methods. **(B)** Representative outputs from the pipeline for samples with positive- and negative- wGI scores.

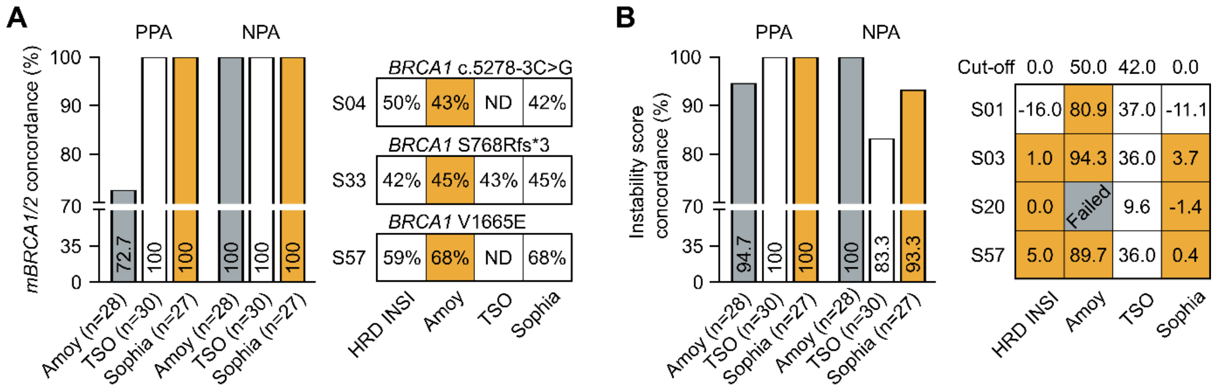


**Figure S3. Analytical validation of the HRD INSIGHT assay.** Negative- and positive-performance agreement (NPA and PPA) of the HRD INSIGHT assay with the HRD Focus Panel (AmoyDx), TruSight Oncology TSO 500 HRD (Illumina) and SOPHiA DDM HRD Solutions (Sophia Genetics) to determine **(A)** *BRCA1/2* mutation status and **(B)** genomic instability status of clinical FFPE ovarian samples. For conflicting samples, the results of individual assays were shown.

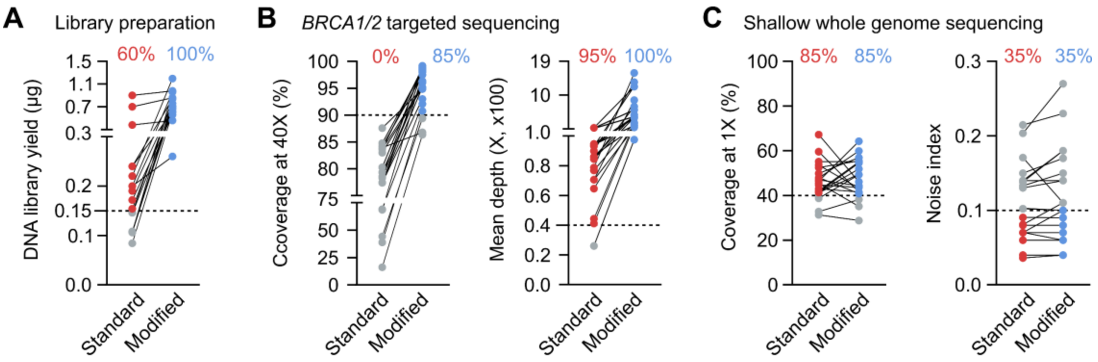


**Figure S4. Modification of wet lab protocol improved library yield and successful rate for *BRCA1/2* targeted sequencing.** A different set of in-house FFPE DNA samples (n = 20) was used to demonstrate the effectiveness of this modification. Using same DNA input, the DNA library yield **(A)** and QC measures for *BRCA1/2* targeted sequencing **(B)** were significantly improved for the modified protocol compared to the standard protocol. **(C)** QC for sWGS data were not different between the two protocols.
